# Analyzing Access to Surgical Services in Central Equatoria State, South Sudan: A Baseline Cross-Sectional Assessment to Inform National Surgical Policy and Planning

**DOI:** 10.64898/2026.04.20.26351353

**Authors:** Majok Deng Akok Deng, Barnabas Tobi Alayande, Ephrem Daniel Sheferaw, Pierrette Ngutete Mukundwa, Tairu Fofanah, Malual Bul Peter, Dario Kuron, Abebe Bekele, Atemthi Dhieu Dau

## Abstract

**Background:** Access to safe, equitable, and affordable surgical and anesthesia care is critical to reducing the burden of surgical diseases in Africa. To understand the state of access in South Sudan, we conducted a baseline assessment of surgical services in Central Equatoria State (CES) in May 2024.

**Objectives:** This study aimed to survey public healthcare facilities in CES capable of providing essential surgical services. We used the capacity to perform cesarean section, laparotomy, and open fracture management—Bellwether procedures—as a proxy for assessing workforce, infrastructure, financing, information management, and service delivery.

**Methods:** We used a validated and contextualized Surgical Assessment Tool developed by the Harvard Program on Global Surgery and Social Change and the World Health Organization. Data were collected at the facility level and summarized descriptively using percentages, means (standard deviations), medians (minimum, maximum), and visualized in graphs, charts, and tables.

**Results:** All three public health facilities assessed could perform Bellwether procedures for their catchment populations. However, workforce availability, financing, and surgical infrastructure were major constraints. The surgical workforce density was 2.27 surgical, anesthesia, and obstetric specialists per 100,000 population. Specialized procedures—such as repair of cleft lip and palate, clubfoot, and hydrocephalus shunt—were unavailable at all sites. None had magnetic resonance imaging (MRI) machines. The total average annual facility budget was $918,850, ranging from $3,960 to $800,000 at the teaching hospital—insufficient for proper operations.

**Conclusion:** While Bellwether procedures are routinely performed, access to quality and affordable care is compromised by deficits in workforce, financing, and infrastructure. We recommend that the Ministry of Health scale this survey nationally and develop a surgical policy and strategic plan focused on improving infrastructure, workforce, and financing for surgical and anesthesia care in South Sudan.

## INTRODUCTION

Surgical conditions contribute to one-third of the global burden of disease (1). Additionally, over 90 percent of the poorest one-third of the global population do not have access to safe, timely, and affordable surgical services (2). In addressing these challenges, surgical infrastructure, financing, information management, surgical governance and service delivery are key pillars that must be developed and strengthened in any country (3). Baselining of surgical systems capacity is a key step in the cycle of planning for national services, and while this has been done in a number of sub-Saharan African countries, has not yet been carried out in South Sudan (4). Surgical capacity in parts of Rwanda, Kenya, Tanzania, Ethiopia, and Malawi have been assessed with Surgical Anaesthesia and Obstetric provider density ranging from 0.5-2.4 per 100,000 population, infrastructure varying widely, and 25-52% of facilities being capable of performing Bellwether procedures (5–9).

The facility readiness for provision of surgical healthcare in South Sudan is unknown. Capacity for service delivery of the ‘Bellwether’ procedures (including cesarean sections, laparotomies, and management of open fractures) at district and tertiary-level facilities is currently undocumented. In addition, information management, surgical financing, and the current state of surgical infrastructure has not yet been described.

Conducting a baseline study to provide a snapshot of where South Sudan lies in terms of access to surgical and anesthesia services is the second step in the development of a national surgical policy and strategic plan (4). Starting this assessment from Central Equatorial State, which hosts the country capital, Juba, will catalyze phased implementation of such studies across the country and provide the much-needed surgical services access data for policy makers, implementers, and other important stakeholders involved in surgical and anesthesia care in Central Equatoria State and South Sudan at large. When such data is made available, gaps in the delivery of quality surgical services in Central Equatoria State (CES), South Sudan, can be identified and addressed.

The aim of this study was to assess the capacity of selected public healthcare facilities in CES, South Sudan to provide surgical healthcare. Specific objectives were to:

1. Assess the availability of surgical infrastructure in three district and tertiary facilities in South Sudan as of May 2024.
2. Assess the surgical service delivery using the proxy of capacity to perform Bellwether procedures (cesarean sections, laparotomies, and management of open fractures) in three district and tertiary facilities in South Sudan as of May 2024.
3. Assess the facility information management, surgical financing, and governance at three district and tertiary facilities in South Sudan as of May 2024.

The overarching objective of this study was to generate baseline data to inform national surgical policy and planning. Using a cross sectional, hospital-based, walk-through survey methodology, we carried out hospital facility assessments in CES, South Sudan focusing on five domains of the surgical healthcare system.

## METHODS

### Study Setting

The study setting was Central Equatoria State (CES), one of the ten states and three administrative areas in South Sudan. Demographic projections by City Population showed that CES has a population of 1.57 million, a land area of 43,033 square kilometers, and a population density of 36.59 people per square kilometer (18). CES was purposely chosen for this assessment because it hosts Juba, the capital city of South Sudan. Conducting the study within this state was intended to reflect the overall maximum access to surgical care services in the country. CES is made up of six counties: Terekeka, Morobo, Yei, Lainya, Kajo-keji and Juba. CES has only two public county (first-level referral) hospitals, and one public teaching hospital for adult populations.

### Study Design

This was a facility-based, cross-sectional study with quantitative methodology.

### Facility selection

Total sampling of county and tertiary public health referral facilities providing care for adult populations in CES, South Sudan, was performed. The list of these facilities was obtained from the Ministry of Health (MOH) record. The highest referral facility for Juba County is Juba Teaching Hospital, Yei and Kajo Keji counties have Yei Civil Hospital and Kajo Keji Hospital, respectively. We excluded Al Sabah children’s hospital, a paediatric tertiary facility, as our focus was on facilities that could provide Bellwether procedures (including caesarean sections) as a measure of service delivery. The recruitment period for the study participants (health facilities and personnel interviewed) was done from 23^rd^ May, 2024 to 31^st^ May, 2024.

### Data collection tools

A validated Surgical Assessment Tool that was developed by the Harvard Program on Global Surgery and Social Change (PGSSC) and the World Health Organization (WHO) was adapted and used at the facility level to collect data on the key pillars of surgical care (19). Adaptation involved changing unfamiliar terminology of ‘district hospital’ to ‘county hospital’, and face validating this with expert reviewers that were familiar with the content and context. The assessment tool consisted of 169 data points, which were grouped according to the WHO health systems strengthening building blocks (20). As there were only three secondary and tertiary facilities in total in CES, and there were no significant changes applied, piloting of the tool was not considered necessary (19).

By using the tool, data on service delivery, infrastructure, workforce, health information management and financing were collected from each selected facility. The variables collected by this tool included the general information about the hospitals, such as its name, address, level, and type of facility being investigated as well as information of health professional who was interviewed or filled the form and various questions about available infrastructure, service delivery, workforce, information management, and financing of the local surgical care services. Data on human resources for health were collected from facility administrators and surgical unit heads. Data on surgical procedures was collected from surgical logbooks, financial data was obtained from facility finance departments, while other data was obtained from relevant departments during a facility walk-through.

### Ethics and Informed Consent

Ethics approval was obtained from the South Sudan Ministry of Health Research Ethics Review Board (MOH/RERB/A-30/2024) and the University of Global Health Equity Institutional Review Board (UGHE-IRB/2024/280). Permission to conduct the study was sought from the CES Ministry of Health and subsequently from the facility heads. This research was conducted according to the principles of the Declaration of Helsinki (21). Written informed consent was obtained from each facility. The study strictly adhered to the ethical guidelines outlined in the Helsinki Declaration for research involving human subjects. Before the hospital facility walkthroughs, the facility administrator provided written informed consent.

### Data collection procedures

Contact with point-persons at the various facilities was made both one week and one day in advance of data collection. The point-person and relevant leadership at the facility were reminded of the purpose of the data collector’s visit one day before data collection. On the day of data collection, information was primarily collected from the hospital directors and triangulated with information from other responsible individuals at appropriate units within the facility. During data collection, data collectors walked through the facility in a systematic manner, visiting relevant departments to ensure accuracy of data collected. Data was collected electronically using the KoboCollect mobile app into a facility characteristics template. Where physical facility walkthrough was not possible due to insecurity, virtual walkthrough using video call was performed.

### Operational Definition

In this study, surgery-capable hospitals by definition refer to facilities that have the capacity to perform at least all three ‘Bellwether’ procedures – caesarean section, laparotomy and management of open fractures (2). The proportion of health facilities capable of doing the Bellwether procedures refers to the number of facilities that offer all three procedures.

### Data analysis

The data collected during this study was quantitatively analyzed and summarized descriptively using percentages, mean (standard deviation), median (minimum, maximum), as well as visually in graphs, charts, and tables. First, the data were downloaded from KoboCollect, cleaned using MS Excel version 2021, and then analyzed with IBM SPSS version 23 software.

## RESULTS

### General Facility Characteristics and Surgical Infrastructure

Of the three facilities surveyed in this study, two were county (first-level referral) hospitals and one was a teaching hospital. All of them are public facilities serving a total population of 1,367,031 people. The highest volume facility, Juba Teaching Hospital, saw 4,820 major surgical patients from June 2023 to May 2024, while Kajo Keji County Hospital, the lowest volume facility, received 450 in the same period. General facility characteristics are shown below Table 1.

**Table 1:** Admissions, outpatients, hospital beds, surgical beds, functional operating rooms, PACU beds, ICU beds and functional ventilators in the ICU at the assessed facilities in CES, South Sudan, May 2024.

| Variables | Median | Minimum | Maximum | Total |
| --- | --- | --- | --- | --- |
| Admissions in a year | 1102.5 | 240.0 | 7284.0 | 17558.0 |
| Outpatients seen in a year | 12960.0 | 480.0 | 34150.0 | 80270.0 |
| Inpatient hospital beds | 37.5 | 13.0 | 300.0 | 665.0 |
| Surgical beds | 9.0 | 4.0 | 97.0 | 110.0 |
| Functioning operating rooms<br>(major and minor) | 3.0 | 3.0 | 4.0 | 10.0 |
| Post-anesthesia care beds (0 for<br>none) | 2.0 | 1.0 | 6.0 | 9.0 |
| Advanced care/ICU beds | 3.0 | 1.0 | 24.0 | 28.0 |
| Functional ventilators in the ICU | 1.0 | 1.0 | 3.0 | 5.0 |

***CES:** Central Equatoria State; **ICU:** Intensive care unit; **PACU:** Post-anesthesia care unit.*

### Service delivery

#### Capacity for ‘Bellwether’ procedures

All three health facilities surveyed had the capacity to offer Bellwether procedures-laparotomy, cesarean section and management of open fractures.

#### Surgical Volume

Surgical volume for surgical procedures across the three facilities was 6,134 cases, representing a surgical volume of 390.7 procedures per 100,000 population.

#### Surgical Safety

Regarding the safety and quality of surgical care, only one facility reported 100% use of the WHO Surgical Safety Checklist all the time, while 100% of the facilities reported 100% use of pulse oximetry in their operating room (OR) all the time, as shown in Table 2.

**Table 2:**
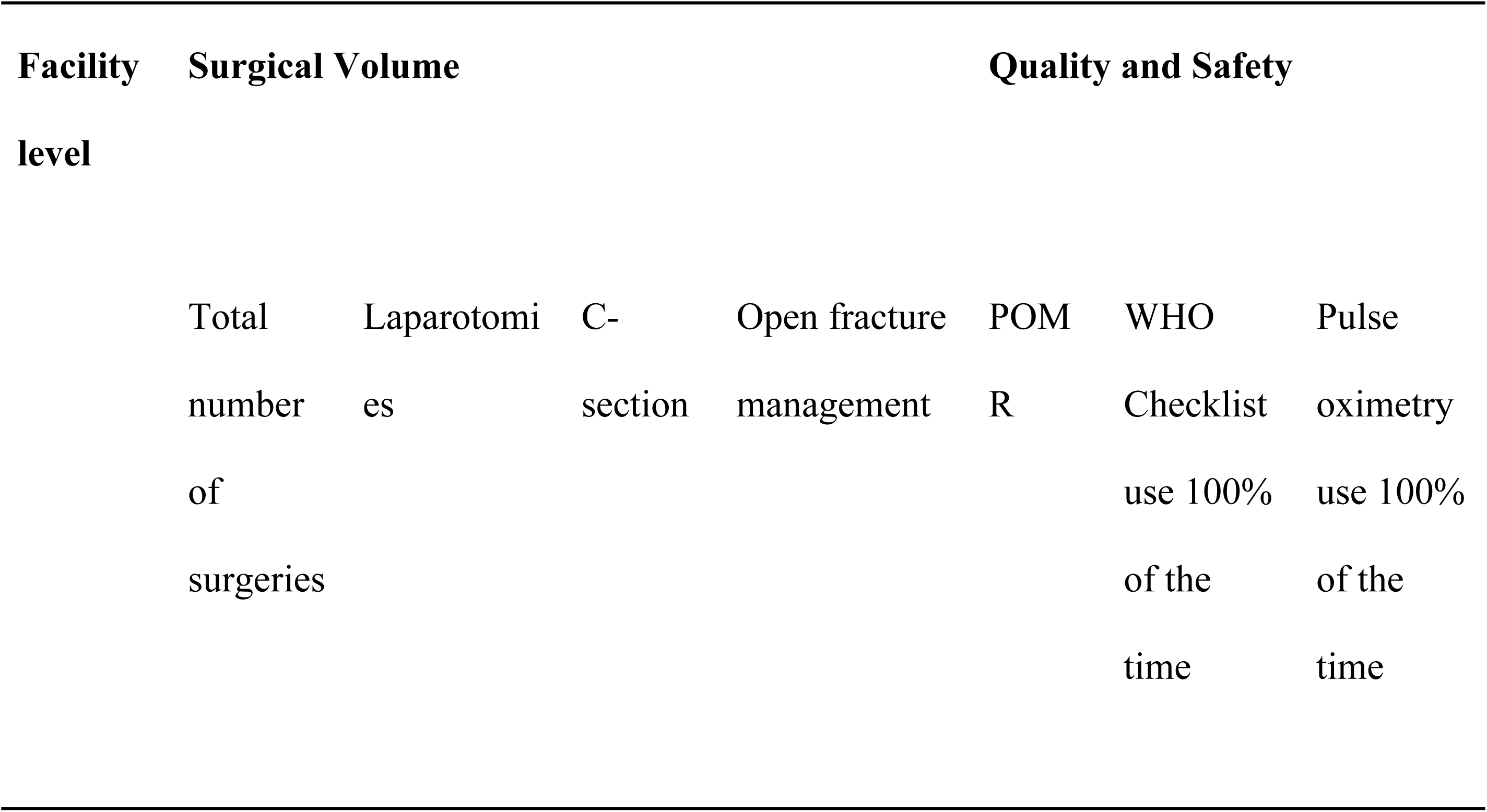
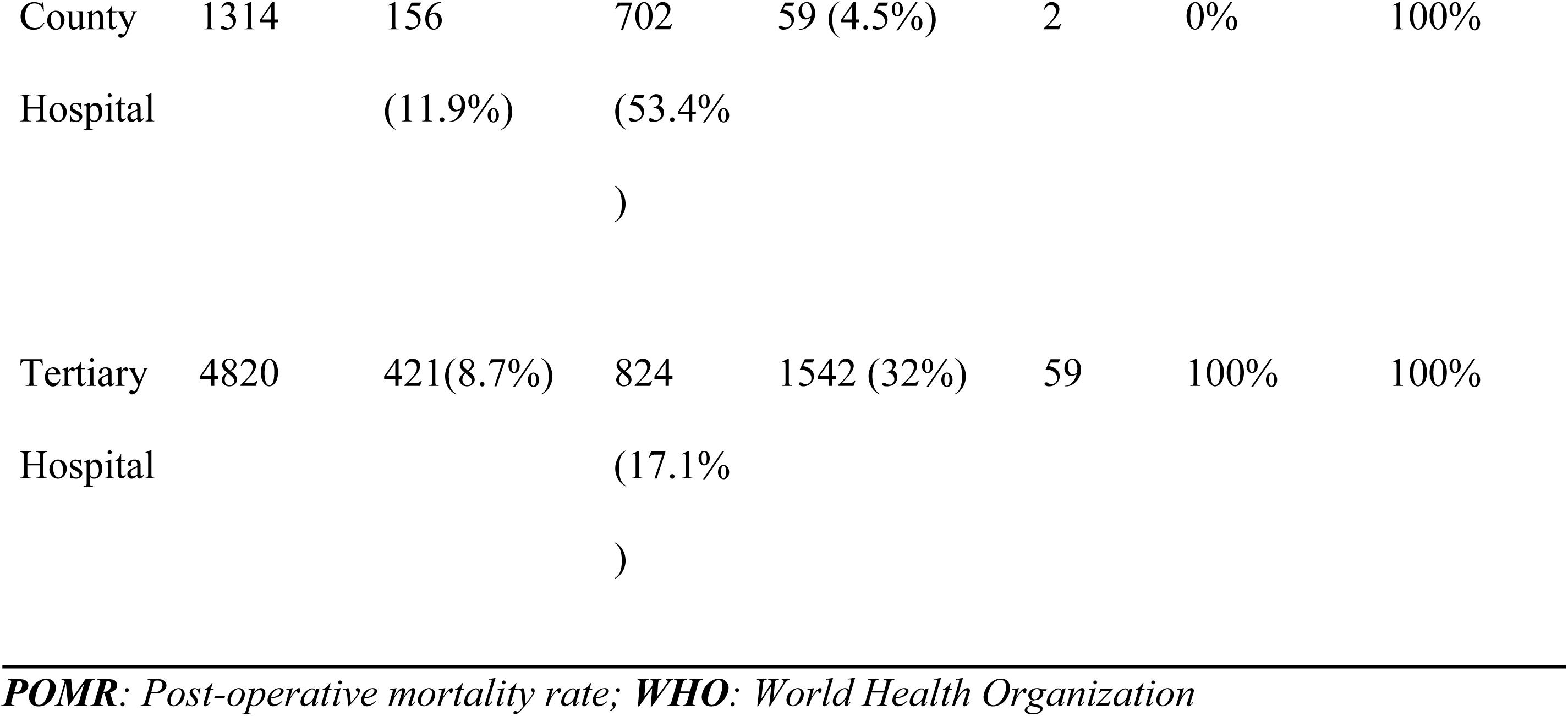
Surgical volume, quality and safety at the assessed health facilities in Central Equatoria State, South Sudan, May 2024.

#### Infrastructure

The availability of oxygen (often available, 100%), electricity (66.7%), and internet facilities (33.3%) was low, while running water was always available at two facilities (66.7%). Radiological infrastructure facilities, including X-ray machines and ultrasound scanners, were almost always available at 100% and 66.7% of facilities, respectively, while a computed tomography (CT scanner was available in only 33.3% of facilities. None of the public health facilities in Central Equatoria State had a magnetic resonance imaging machine. Laboratory capacity for complete blood count test was available only at 33.3% of hospitals, while urinalysis and infectious disease panel were available in half of the facilities Figure 1.

**Figure 1:**
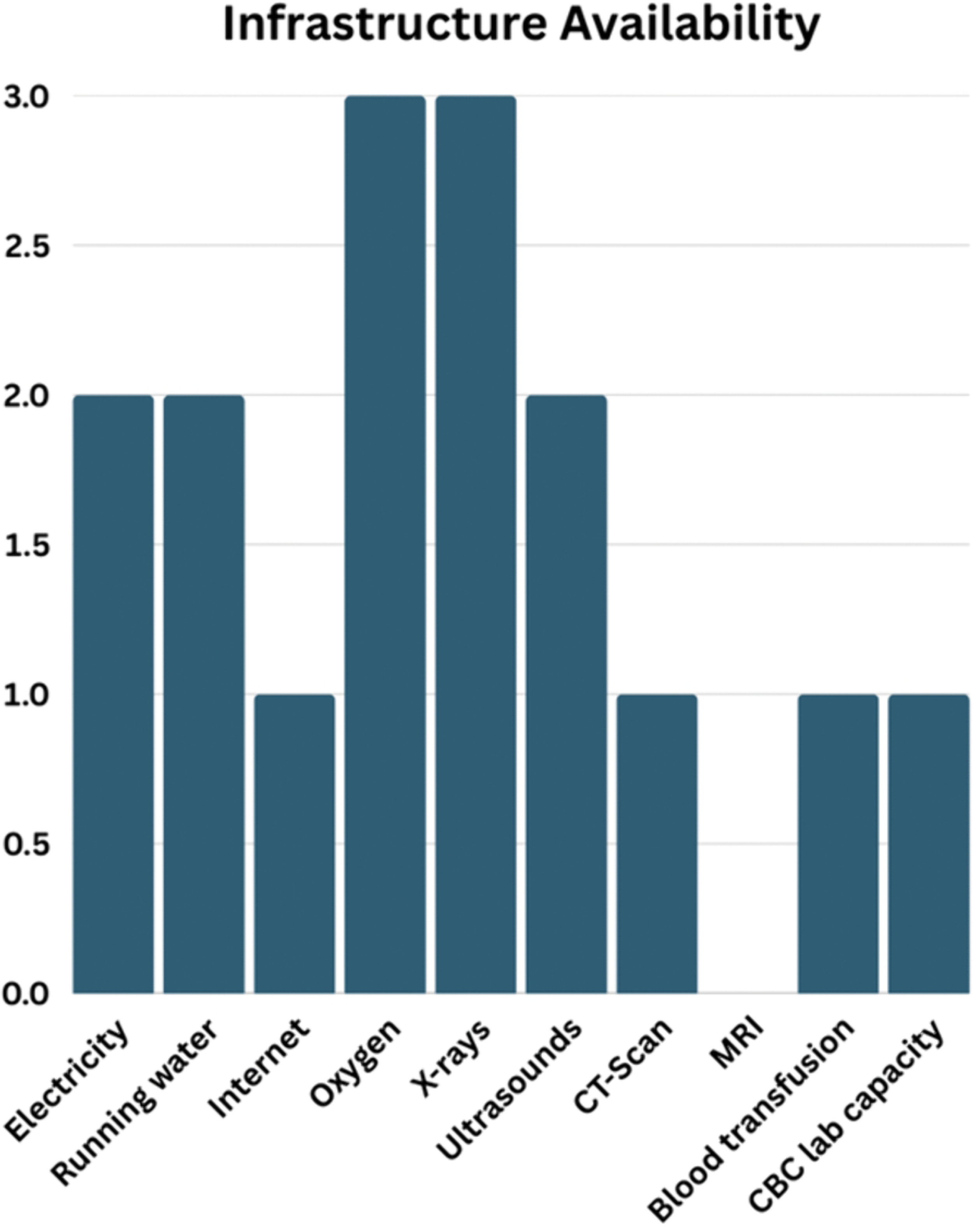
Utility, blood, radiology and laboratory tests availability at the assessed facilities in CES, South Sudan, May 2024.

Regarding blood supply, only one facility (33.3%) reported almost always having the capacity to administer blood transfusion within two hours and another two facilities without such a capacity (66.7%) as shown in Figure 2.

**Figure 2:**
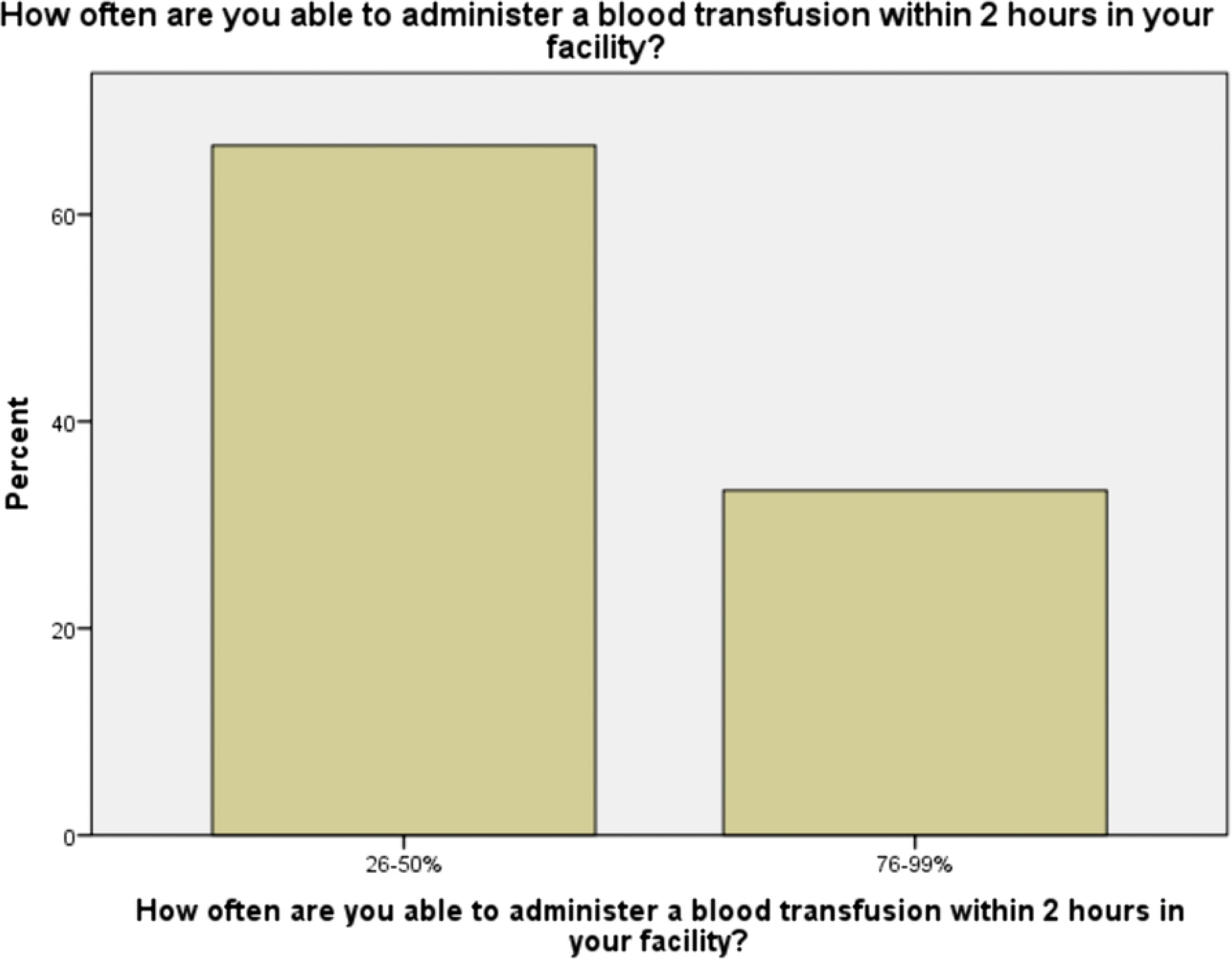
Administration of blood transfusion within 2 hours at the facility in CES, South Sudan, May 2024

#### Workforce

The total number of surgical, anesthetic and obstetric (SAO) specialists across the facilities were 31 in number, but they were more concentrated at the tertiary hospital (87.1%). There were 55 general doctors providing surgical and anesthesia care at these surgery-capable health facilities. Task-shifting was also reported with 36 non-physicians providing the surgical and anesthesia care. The workforce densities for midwives and surgical nurses were higher than the other surgical care cadres with the former being fairly distributed among the different levels of facilities. However, the supporting workforce availability (such as radiologists, biomedical technicians, pathologists) was limited and the few available were mostly found at the tertiary hospital in the capital city as shown in Figure 3.

**Figure 3:**
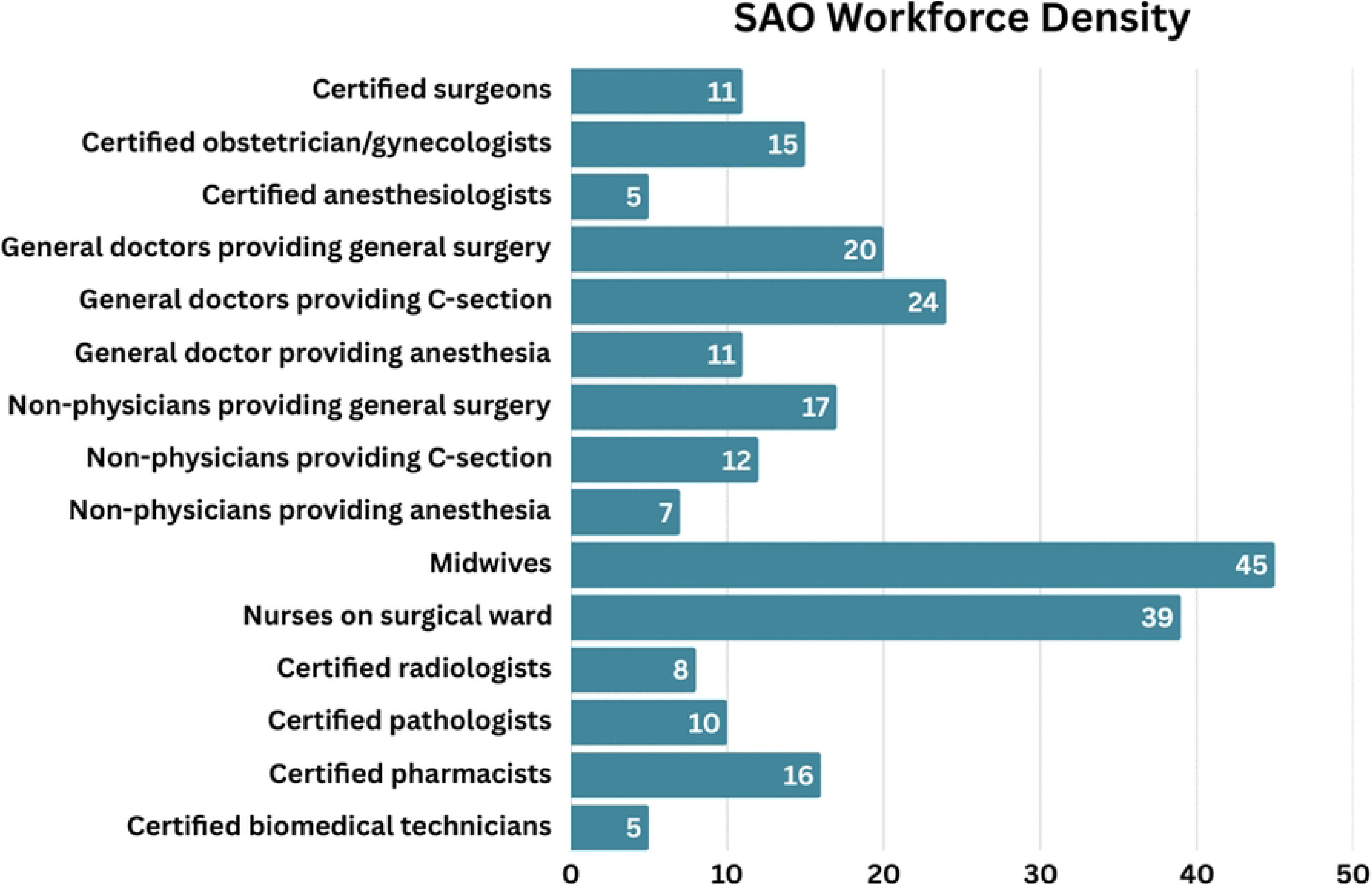
Surgical, obstetric and anesthesia (SOA) workforce at assessed facilities in CES, South Sudan, May 2024.

The SAO specialists’ density per 100,000 at the assessed facilities was 1.97 SAO specialists per 100, 000 population. The non-specialist providers’ density per 100,000 at the assessed facilities was 5.80 non-specialist providers per 100,000 population. The most common and underrepresented SAO specialist groups were the obstetrician-gynecologists and anesthesiologists accounting for 48.4% and 16.1% of the SAO specialists respectively.

#### Service Delivery

The most and least commonly performed obstetric and gynecologic procedures at the assessed hospitals (first-level referral) were cesarean section and vasectomy with median (min, max) of 245.5 (239, 528) and 0.0 (0.0, 0.0) cases respectively. Of the general surgical procedures, the most and least performed procedures were appendectomy and colostomy with median (min, max) of 35 (0, 280) and 0 (0, 5) cases respectively. The most performed traumatic injury and non-traumatic orthopedic procedures were fracture reduction and debridement of osteomyelitis with median (min, max) of 24 (0, 50) and 3 (0, 105) procedures respectively. Repair of cleft lip and palate, clubfoot, shunt for hydrocephalus, cataract extraction or insertion of intraocular lens and eyelid surgery for trachoma were not found at all of the assessed facilities as shown in Table 3.

**Table 3:**
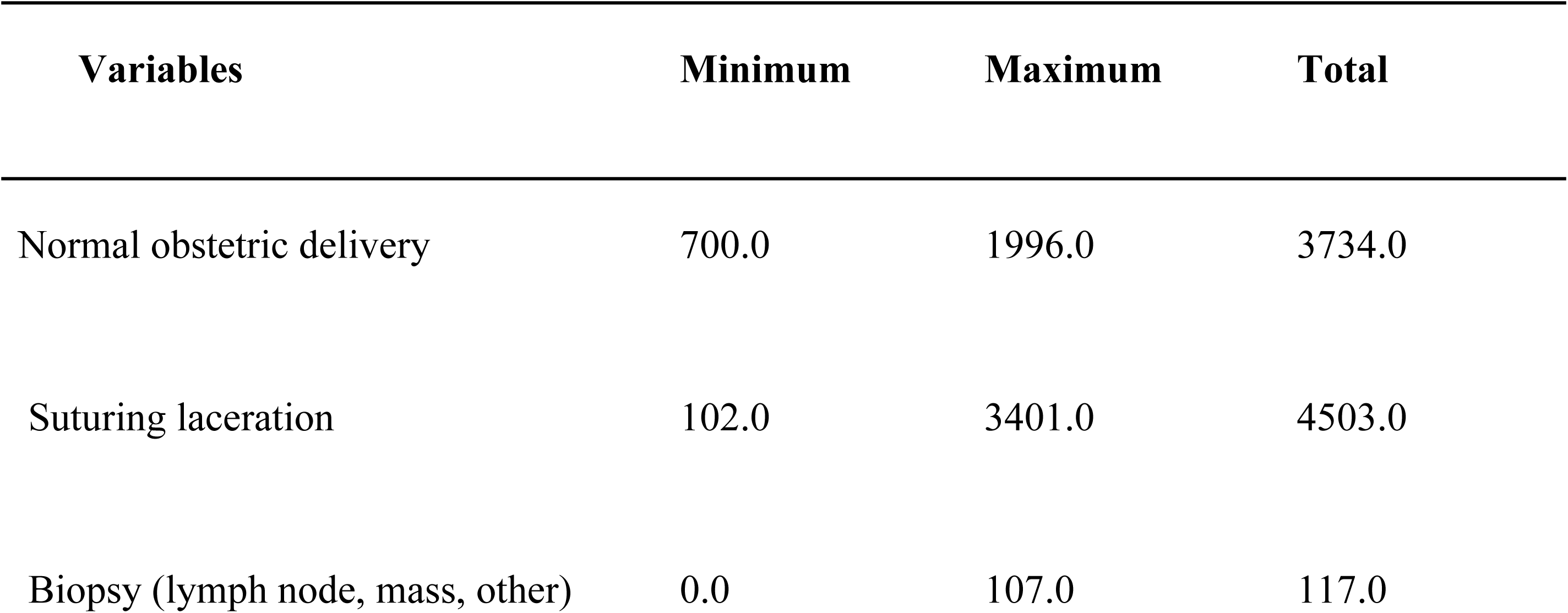
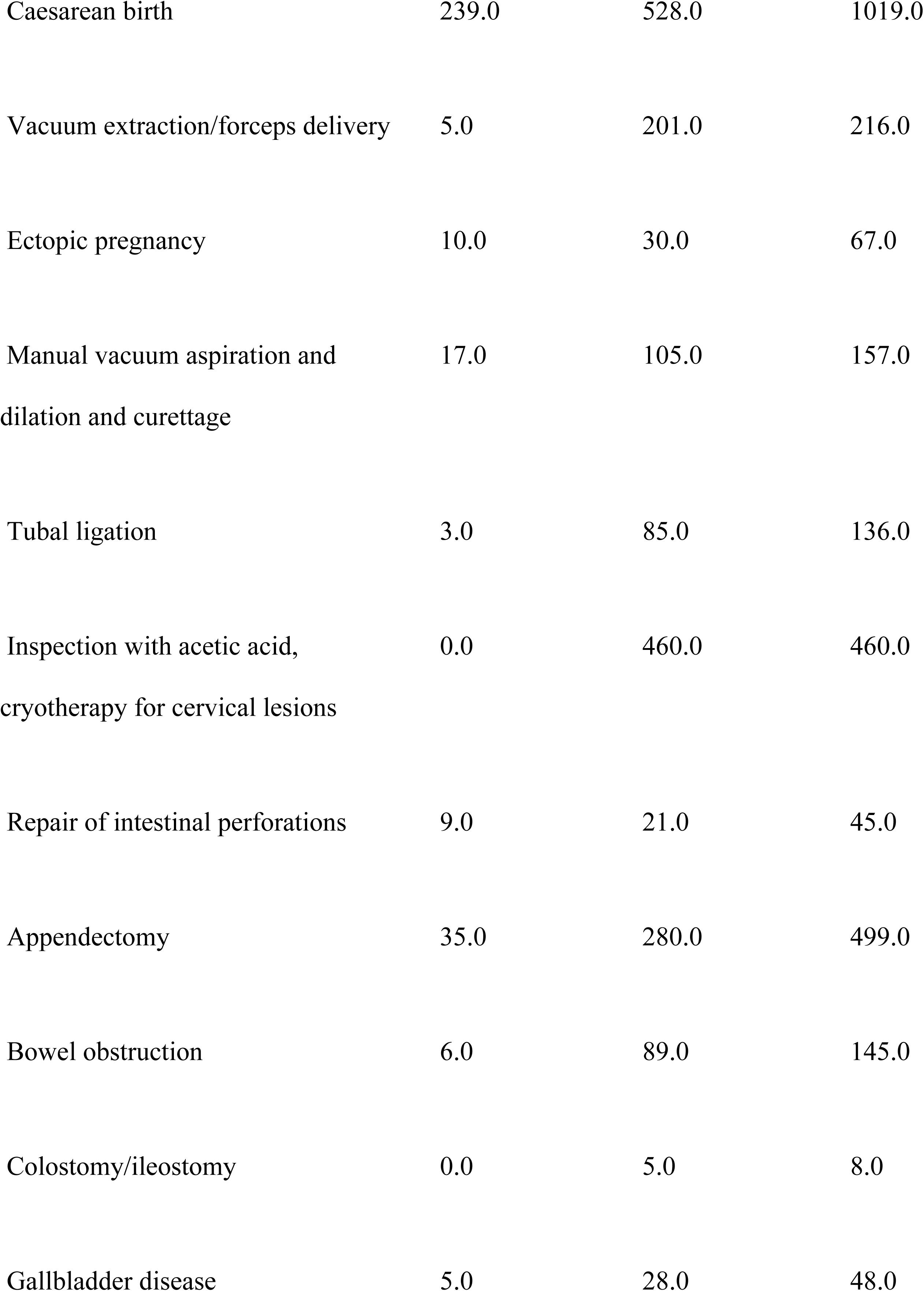
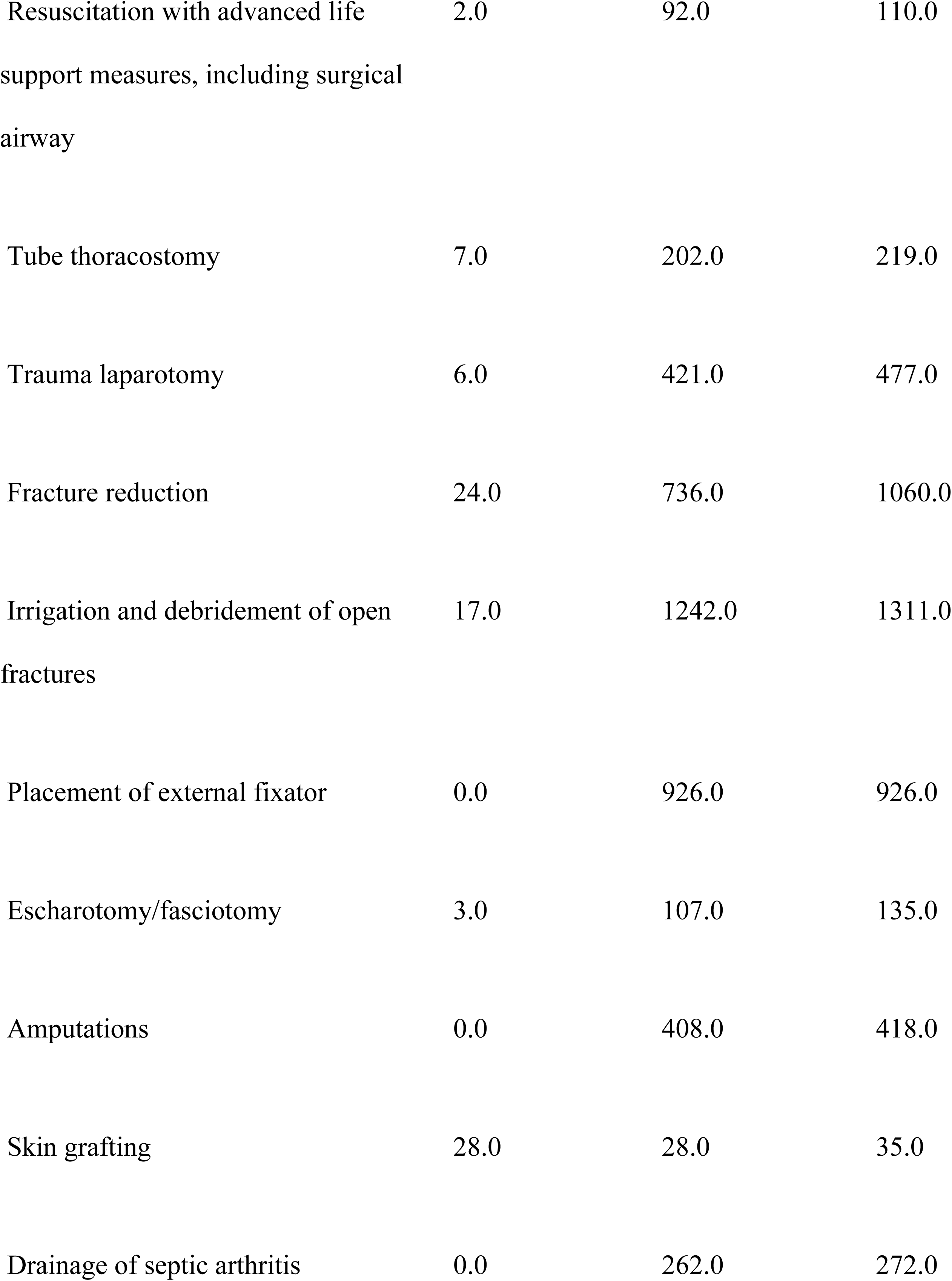
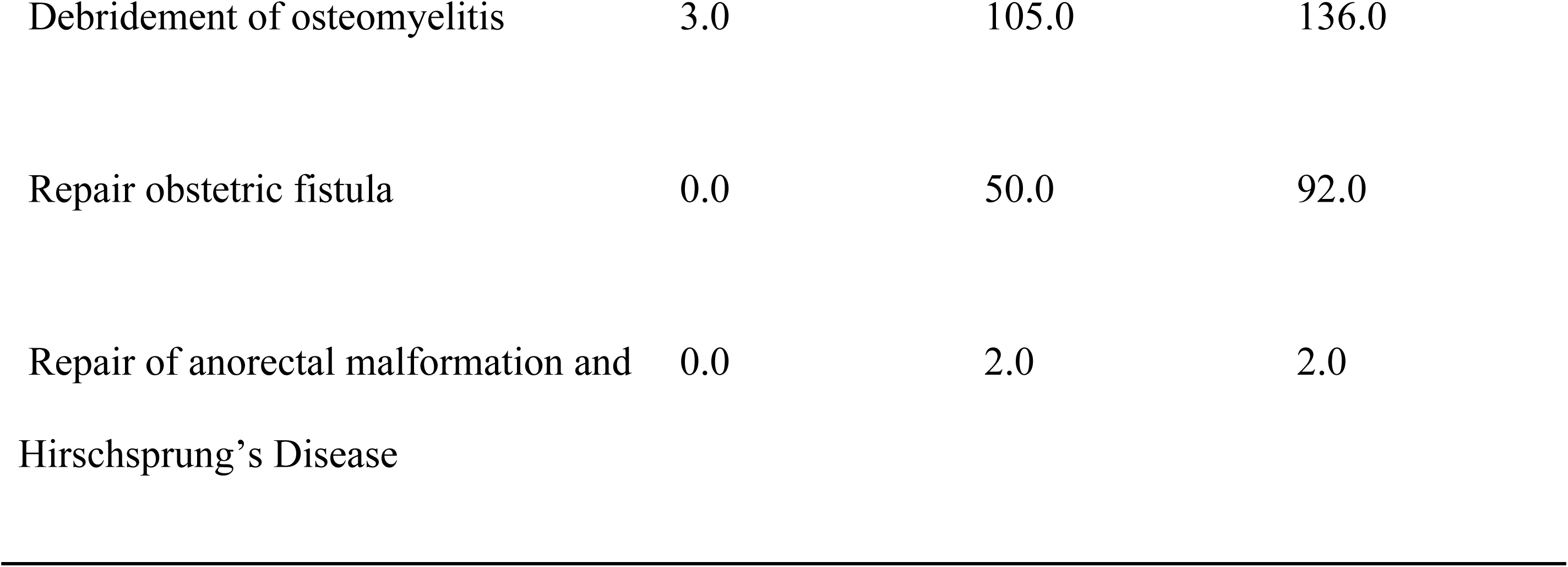
Surgical procedures available at the assessed facilities in Central Equatoria State, South Sudan, May 2024.

The infrastructure to deliver surgical and anesthesia services were generally inadequate at all levels of facilities. Only few supplies and equipment were always available 100% of the times as shown in Table 4.

**Table 4:** Pharmacy, surgical and anesthesia equipment and supplies that were almost always available at the assessed facilities in Central Equatoria State, South Sudan, May 2024. N=3.

| Pharmacy | Facilities | Anesthesia | Facilities | Surgical | Facilities |
| --- | --- | --- | --- | --- | --- |
|  | with | Equipment & | with | Equipment & | with |
|  | equipment | Supplies | equipment | Supplies | equipment |
|  | almost |  | t almost |  | t almost |
|  | always |  | always |  | always |
|  | available |  | available |  | available |
|  | n (%) |  | n (%) |  | n (%) |
| Inhalational<br>general anesthesia | 1(33.3) | Adult<br>oropharyngeal<br>airway | 0(0) | Electrocautery | 0(0) |
| IV sedation<br>anesthesia | 2(66.7) | Pediatric<br>oropharyngeal<br>airway | 1(33.3) | Autoclave<br>sterilizer | 1(66.7) |
| Spinal anesthesia | 3(100) | Adult<br>endotracheal<br>tube | 1(33.3) | Forceps | 1(33.3) |
| Regional<br>anesthesia | 2(66.7) | Pediatric<br>endotracheal<br>tube | 1(33.3) | Syringes with<br>needles | 2(66.7) |
| Perioperative<br>antibiotics | 2(66.7) | Adult<br>laryngoscope | 1(33.3) | Scalpel | 2(66.7) |
| IV fluids | 2(66.7) | Pediatric<br>laryngoscope | 1(33.3) | Scissors | 1(33.3) |
| Muscle<br>relaxants/paralyti<br>cs | 2(66.7) | Adult facemask<br>bag valve | 1(33.3) | Needle holder | (66.7) |
| Sedatives | 3(100) | Pediatric<br>facemask bag<br>valve | 1(33.3) | Retractor | 2(66.7) |
| Vasopressors | 0(0) | Difficult airway<br>kit (LMA) | 1(33.3) | Sterile gloves | 2(66.7) |
| Postoperative<br>narcotics | 1(33.3) | Adult Magill<br>forceps | 0(0) | Urinary<br>catheters | 1(33.3) |
|  |  | Pulse oximeter | 2(66.7) | Gowns | 2(66.7) |
|  |  | Stethoscope | 2(66.7) | Disinfectant<br>hand wash | 2(66.7) |
|  |  | Suction<br>apparatus | 1(33.3) | Sterilizing skin<br>prep | 2(66.7) |

#### Information Management

The study revealed that all facilities have personnel responsible for managing medical records, with 33.3% of the facilities utilizing both paper-based and computer-based systems and 66.7% using a paper-based system. 66.7% of the facilities have patients’ charts available at multiple visits. Only two facilities reported using telemedicine, such as remote assistance during surgery, in their surgical care. Research-wise, none of the facilities reported any ongoing facility-level research or surgical department research projects.

Table 5 shows findings from prospective data collection, postoperative data collection for certain conditions, and the frequency of reports to the health authority, as well as quality improvement projects.

**Table 5:**
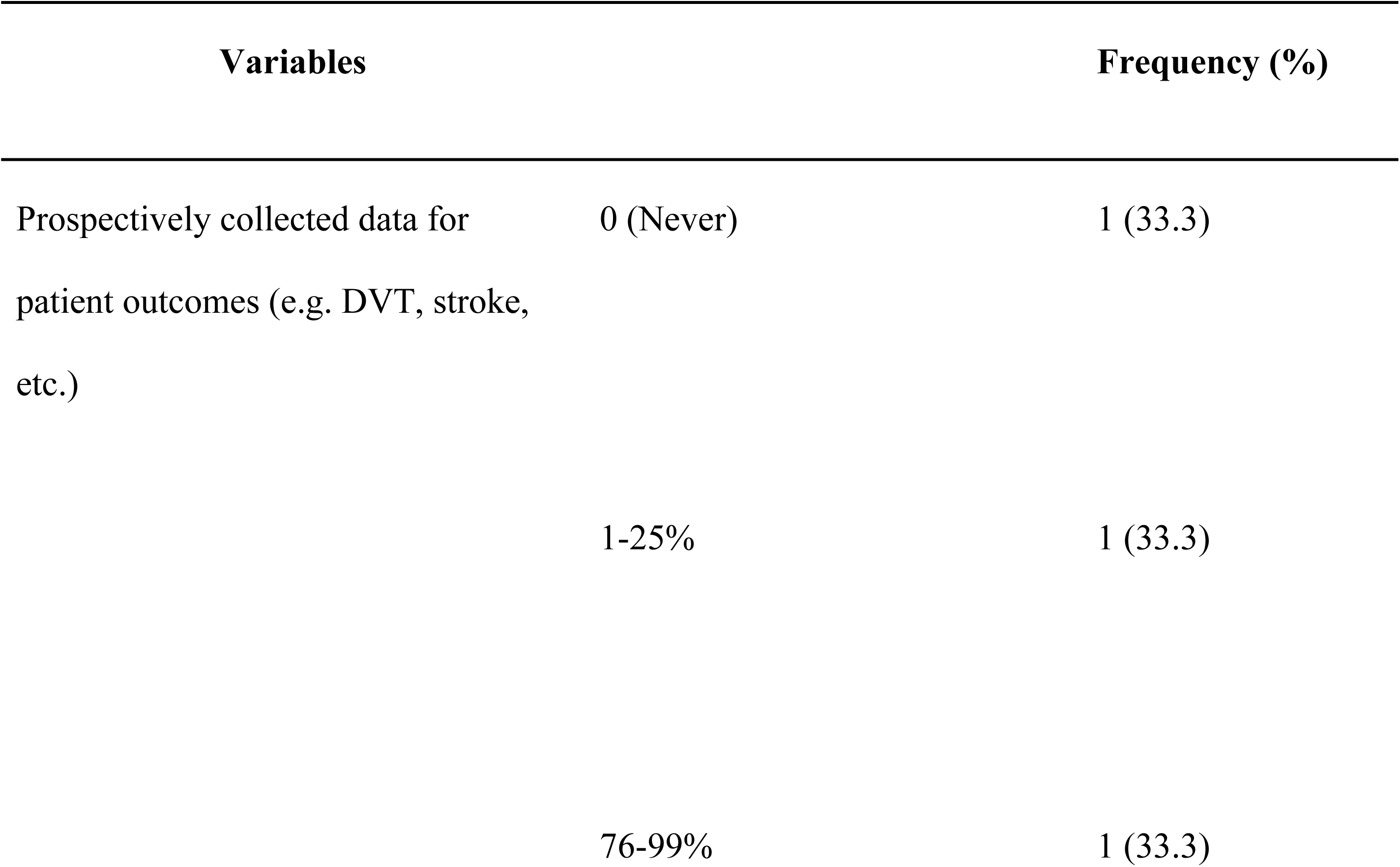
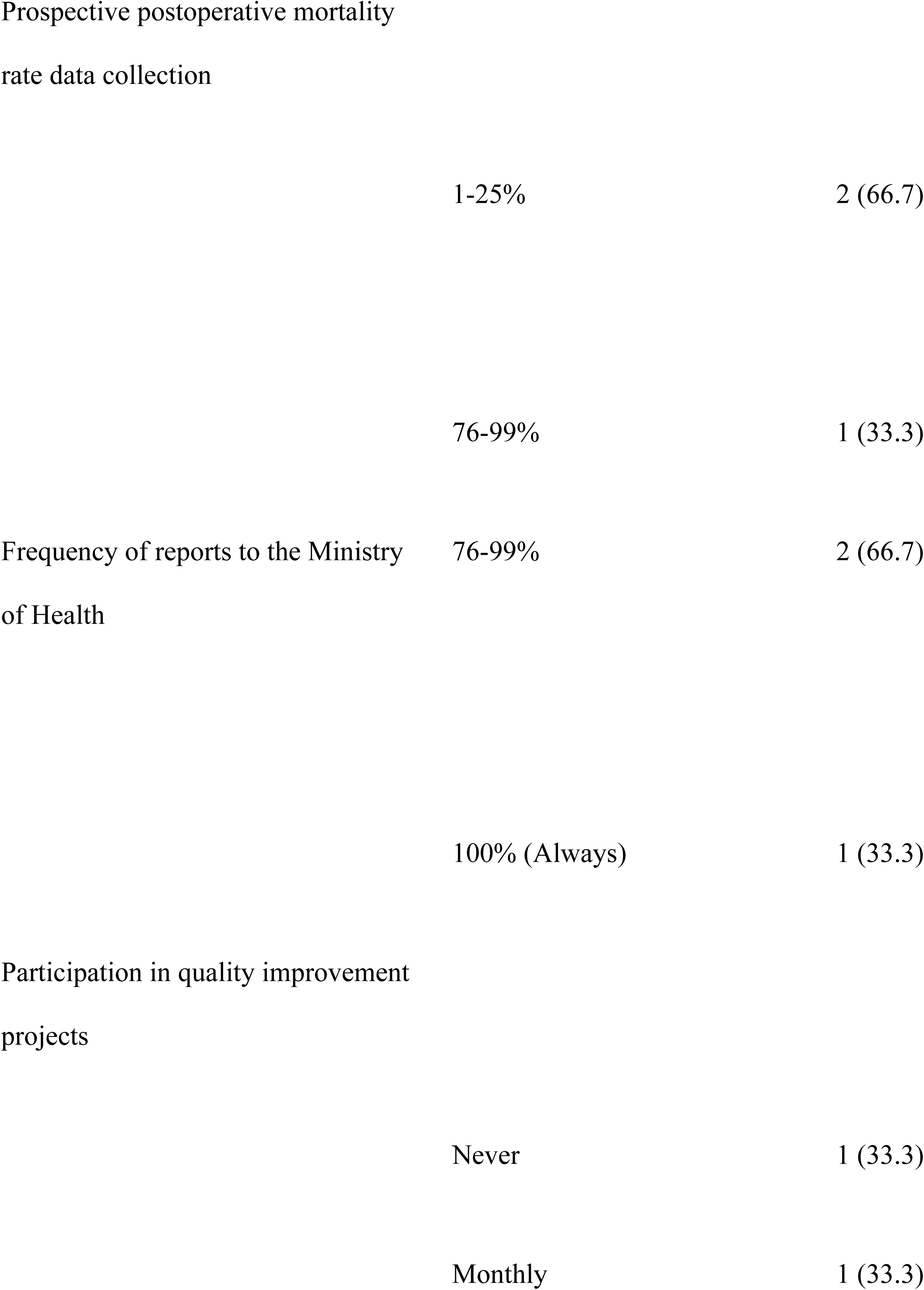
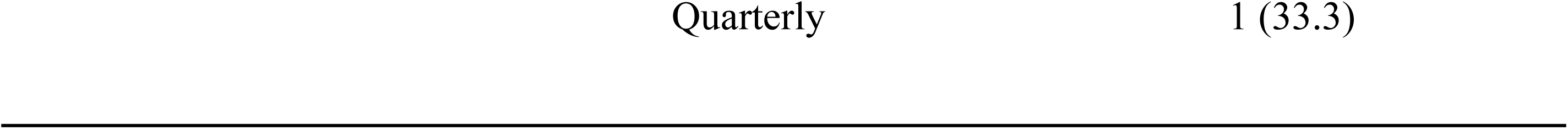
Facility information management and research capacity at the assessed facilities in CES, South Sudan, May 2024.

#### Health Financing

The total average annual facility budget for the facilities was $ 269,616.67, ranging from $3,960 to $800,000 at the teaching hospital. Only one of the facilities reported having 100% insurance coverage for its patients; another facility reported allocating between 1% and 25% of its facility budget to surgery and anesthesia care, with the remaining facility having no insurance coverage for its patients. The highest out-of-pocket costs for the patients were from surgery-associated medication and transportation per surgery, as shown in Table 6.

**Table 6:**
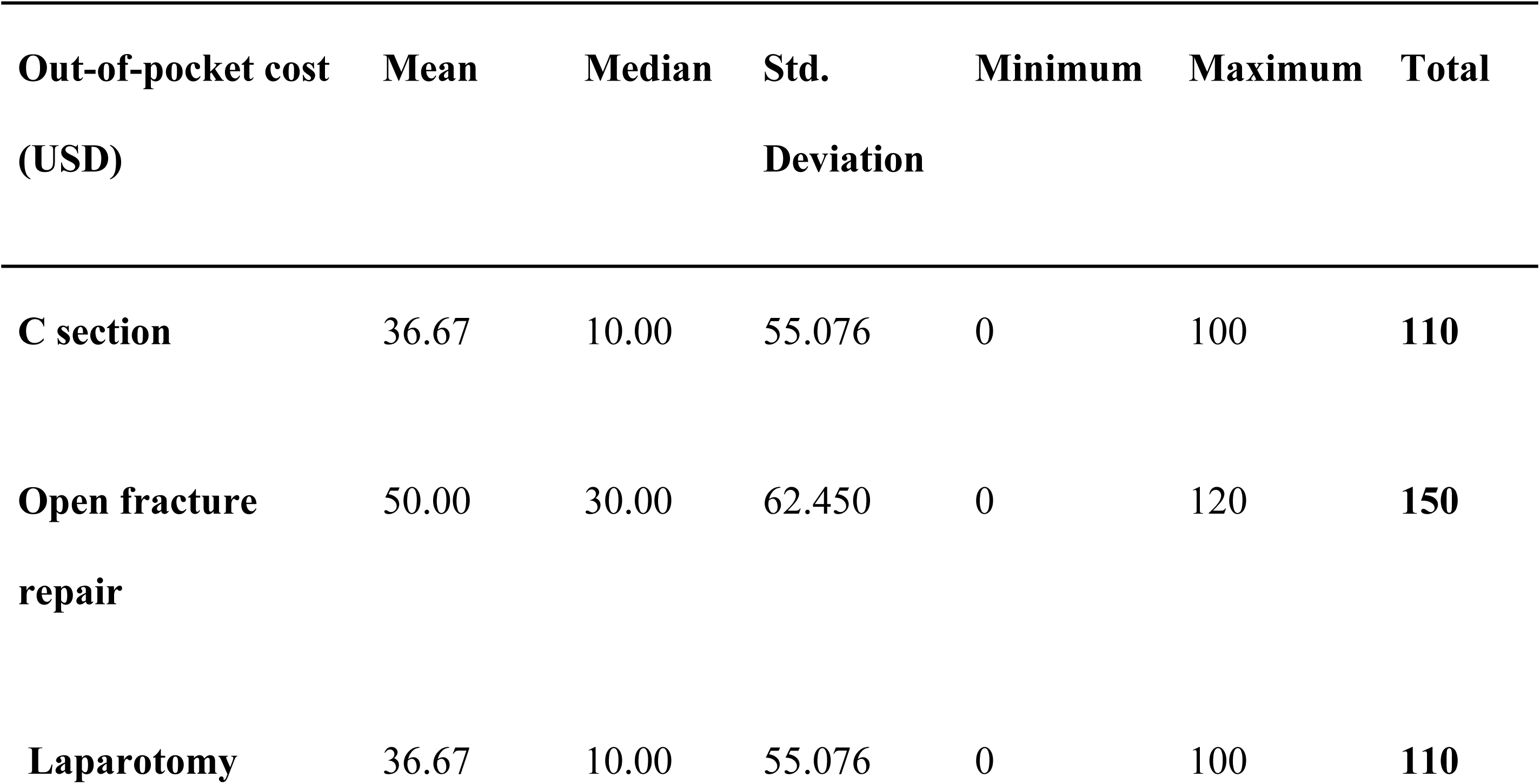
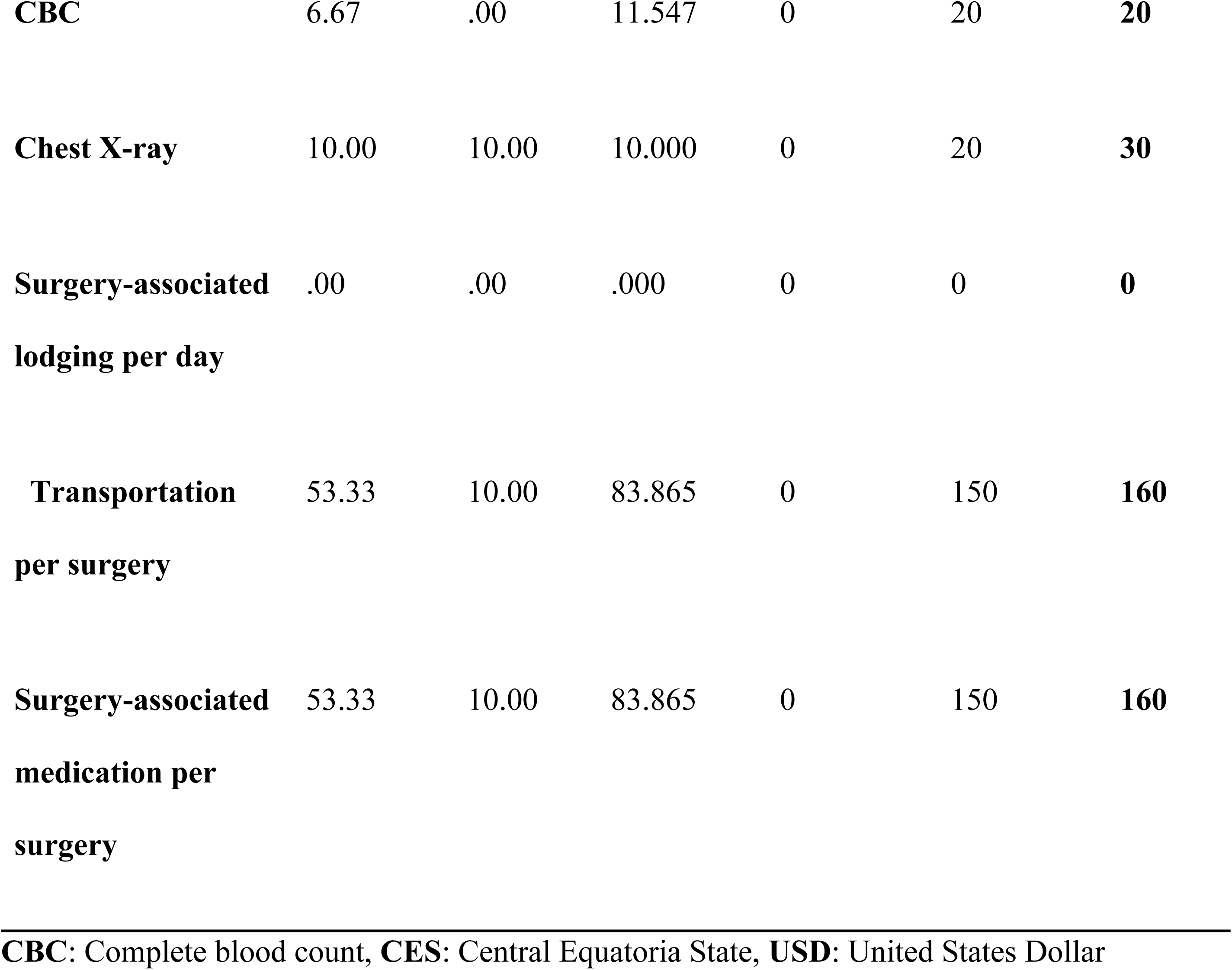
Patients’ out-of-pocket costs for medical and non-medical costs associated with surgery at assessed facilities in CES, South Sudan, May 2024.

| Out-of-pocket cost<br>(USD) | Mean | Median | Std.<br>Deviation | Minimum | Maximum | Total |
| --- | --- | --- | --- | --- | --- | --- |
| **C section** | 36.67 | 10.00 | 55.076 | 0 | 100 | **110** |
| **Open fracture**<br>**repair** | 50.00 | 30.00 | 62.450 | 0 | 120 | **150** |
| **Laparotomy** | 36.67 | 10.00 | 55.076 | 0 | 100 | **110** |
| **CBC** | 6.67 | .00 | 11.547 | 0 | 20 | **20** |
| **Chest X-ray** | 10.00 | 10.00 | 10.000 | 0 | 20 | **30** |
| **Surgery-associated**<br>**lodging per day** | .00 | .00 | .000 | 0 | 0 | **0** |
| **Transportation**<br>**per surgery** | 53.33 | 10.00 | 83.865 | 0 | 150 | **160** |
| **Surgery-associated**<br>**medication per**<br>**surgery** | 53.33 | 10.00 | 83.865 | 0 | 150 | **160** |

266 **CBC:** Complete blood count, **CES:** Central Equatoria State, **USD:** United States Dollar

## DISCUSSION

This baseline assessment provides an essential overview of the surgical care landscape in Central Equatoria State (CES), South Sudan, using Bellwether procedures as a proxy of access and capacity. While all three assessed public facilities were capable of performing cesarean sections, laparotomies, and open fracture management, significant deficits were observed across multiple domains of surgical care, with major gaps in infrastructure, workforce, financing, and service delivery.

The finding that all surveyed facilities could offer Bellwether procedures aligns with the minimum standard recommended by the Lancet Commission on Global Surgery (LCoGS), which identifies these procedures as essential indicators of a functioning surgical system (2). However, this does not imply that surgical services in CES are adequate or equitably distributed. Notably, only the tertiary-level facility, Juba Teaching Hospital, was able to provide a broader range of procedures, including higher surgical volumes and postoperative care, while county hospitals remained under-resourced.

This finding agrees with the broader continental trends. Bekele et al. (2023) report that although some African countries have begun developing NSOAPs, most still face challenges in implementing equitable surgical services due to funding and human resource constraints (4). Similarly, Reddy et al. (2021) highlight systemic barriers such as fragmented governance, donor-driven priorities, and limited technical capacity, which inhibit the development of sustainable surgical systems across sub-Saharan Africa (5).

One of the most crucial findings of this study is the critically low SAO workforce density—2.27 per 100,000 people—far below the LCoGS benchmark of 20 per 100,000 (2). This is consistent with IndexMundi data on South Sudan’s SAO workforce, which also reports a ratio of 0.32 per 100,000 nationally (10). This inadequacy contributes to task shifting and sharing, a practice also observed in Aweil Civil Hospital, where non-physician clinicians provide essential surgical services (14). While task sharing has been advocated as a short-term strategy in resource-poor settings, its long-term safety and sustainability remain concerns unless accompanied by adequate supervision and standardized training programs (14,19).

Infrastructure limitations further compound access challenges. The inconsistent availability of basic utilities such as electricity (available 66.7% of the time), oxygen, and internet connectivity severely undermines surgical readiness. Diagnostic capacity was also weak; for instance, only one facility had a CT scanner and none had an MRI machine, echoing findings from Bruno et al. (2017) in Madagascar, who emphasized that reliable infrastructure is foundational to safe surgery (19). The limited laboratory capacity (e.g., only one facility consistently capable of conducting complete blood counts) also compromises preoperative risk assessment and postoperative monitoring.

Furthermore, health financing emerged as another critical barrier. Out-of-pocket expenditures for essential procedures like laparotomy and cesarean section remained high in all facilities. This finding supports broader global evidence indicating that lack of financial protection deters timely surgical care and exacerbates health inequities (2,3). In South Sudan, where the health sector receives less than 3% of the national budget (13, 15), systemic underfunding has crippled surgical service delivery and stifled the development of sustainable financing mechanisms, including insurance coverage and strategic purchasing.

The information management systems in the assessed facilities remain weak. Most relied on paper-based records, with no consistent prospective data collection on outcomes or mortality. Moreover, only one facility engaged in any quality improvement initiative. As highlighted by Meara et al. (2015) and Price et al. (2015), the lack of routine data systems and audit mechanisms severely limits accountability, learning, and performance improvement (2,3). Finally, the study’s finding that many essential surgical procedures—such as cleft lip/palate repair, hydrocephalus shunt, and cataract surgery—were not performed at any of the assessed facilities uncovers a significant gap in specialized surgical services. These facilities failed to achieve the packages and procedures for their respective platforms according to the findings from the Disease Control Priorities Project (DCP-3), which notes that while basic emergency surgical services should be available at the district level, more advanced procedures often remain inaccessible in LMICs due to human resource and equipment shortages (22).

In summary, the findings of this study highlight the urgent need for coordinated national policy efforts to improve surgical care access in South Sudan. While Central Equatoria State has relatively better infrastructure and access compared to more remote states, it still falls short of international benchmarks. A nationwide scale-up of similar assessments is necessary to build an evidence base for a national surgical, obstetric, and anesthesia plan (NSOAP), as endorsed by the World Health Assembly Resolution WHA68.15 (3, 4).

### Limitations of the study

This study has several limitations. First, data collection relied partly on retrospective records and self-reports from healthcare workers, which are prone to recall and reporting bias. Although data collectors cross-verified responses through walk-throughs and video calls, subjective inconsistencies may persist.

Second, security challenges in Kajo Keji County restricted in-person data collection, necessitating a virtual walkthrough. Although the virtual approach is validated and has been used in similar contexts, it may have limited the ability to observe contextual factors and infrastructure issues as comprehensively as in-person assessments (20).

Third, the study was geographically limited to one state—Central Equatoria State—which may not be representative of the broader national landscape, especially more remote or underserved regions. CES was purposely chosen due to its relatively better access to surgical care, suggesting that other states may exhibit even more significant deficiencies.

Lastly, the study employed only descriptive statistical methods. While useful for mapping availability and capacity, it does not explore associations or causal relationships between facility characteristics and surgical outcomes. Future research should integrate inferential statistics to provide deeper insights into systemic barriers and enablers.

## CONCLUSION

This baseline assessment reveals a fragile surgical care system in Central Equatoria State. While all assessed public hospitals can provide Bellwether procedures, their capacity to deliver comprehensive and safe surgical care is severely limited by critical shortages in infrastructure, skilled workforce, equipment, and financial protection for patients. The study reveals the urgent need for systemic reforms, starting with the development of a national surgical, obstetric, and anesthesia plan (NSOAP) tailored to the South Sudanese context.

Despite being the host of the national capital, CES exemplifies the broader national challenges—fragmented service delivery, low SAO workforce density, insufficient financing, and inadequate health information systems. Addressing these issues is essential not only for improving surgical outcomes but also for reducing maternal and trauma-related mortality, achieving universal health coverage, and meeting Sustainable Development Goals (SDGs).

### RECOMMENDATION

The Ministry of Health should urgently establish a comprehensive NSOAP to guide investment in infrastructure, workforce development, financing, and service delivery. This strategic plan should align with the Lancet Commission on Global Surgery and WHO recommendations to build surgical systems as a core component of universal health coverage.

County hospitals require targeted upgrades to meet minimum standards for essential surgical care. This includes reliable electricity, oxygen supply, operating rooms, post-anesthesia care units, and basic diagnostic equipment. Strengthening these facilities will help decentralize access to Bellwether procedures and reduce referral delays.

Given the low SAO workforce density, structured task-sharing programs with non-physician clinicians should be scaled up under national guidelines. These programs must include competency-based training, supervision, and certification to ensure safety and quality of care.

The surgical and anesthesia equipment and essential supplies were inconsistently available across all facilities. The Ministry of Health should establish dedicated procurement systems and logistics support to ensure consistent availability of surgical tools, medications, and perioperative resources.

All facilities should adopt standardized data collection tools for surgical outcomes, mortality, and complications. Introducing electronic record systems and supporting regular quality improvement activities—such as morbidity and mortality reviews—will enable better planning, accountability, and learning.

## Data Availability

All relevant data are within the manuscript and its Supporting Information files.

## REFERENCES

1. Shrime MG, Bickler SW, Alkire BC, Mock C. Global burden of surgical disease: an estimation from the provider perspective. Lancet Glob Health. 2015;3:S8–9.

2. Meara JG, Leather AJM, Hagander L, Alkire BC, Alonso N, Ameh EA, et al. Global Surgery 2030: evidence and solutions for achieving health, welfare, and economic development. The Lancet. 2015;386(9993):569–624.

3. Price R, Makasa E, Hollands M. World Health Assembly Resolution WHA68.15: “Strengthening Emergency and Essential Surgical Care and Anesthesia as a Component of Universal Health Coverage”. World J Surg. 2015;39(9):2115–25.

4. Bekele A, Alayande BT, Powell BL, Obi N, Seyi-Olajide JO, Riviello RR, et al. National Surgical Healthcare Policy Development and Implementation: Where do We Stand in Africa? World J Surg. 2023;47(12):3020–9.

5. Shiferaw TB, Gashaw M, Tolera S, Gebremedhin S. Assessing Ethiopia’s surgical capacity in light of global surgery 2030 initiatives: Is there progress in the past decade? Heliyon. 2024;10(7):e28322.

6. Oduor PR, Mbaki MB, Macharia WM. A Narrative Review of Kenya’s Surgical Capacity Using the Lancet Commission on Global Surgery’s Indicator Framework. Glob Health Sci Pract. 2022;10(1):e2100500.

7. Citron I, Jumbam DT, Ulisubisya M, et al. Towards equitable surgical systems: development and outcomes of a national surgical, obstetric and anaesthesia plan in Tanzania. BMJ Glob Health. 2019;4(2):e001282.

8. Gajewski J, Borgstein E, Bijlmakers L, et al. Surgical and anaesthetic capacity of hospitals in Malawi: key insights. Health Policy Plan. 2017;32(7):947–954.

9. Petroze RT, Nzayisenga A, Russoniello N, Ntakiyiruta G, Calland JF. Service delivery, infrastructure, and workforce: a multi-site assessment of surgical capacity in Rwanda. World J Surg. 2012;36(5):1057–1065.

10. Indexmundi. South Sudan - Specialist surgical workforce (per 100,000 population) [Internet]. 2023 [cited 2023 Nov 12]. Available from: https://www.indexmundi.com/facts/south-sudan/indicator/SH.MED.SAOP.P5

11. Alobo G, Reverzani C, Sarno L, Giordani B, Greco L. Estimating the Risk of Maternal Death at Admission: A Predictive Model from a 5-Year Case Reference Study in Northern Uganda. Obstet Gynecol Int. 2022;2022:8947917.

12. Musarandega R, Nyakura M, Machekano R, Pattinson R, Munjanja SP. Causes of maternal mortality in Sub-Saharan Africa: A systematic review of studies published from 2015 to 2020. J Glob Health. 2021;11:04048.

13. World Health Organization. Investigating Maternal Mortality to Save Lives in South Sudan - South Sudan | ReliefWeb [Internet]. 2023 [cited 2023 Nov 12]. Available from: https://reliefweb.int/report/south-sudan/investigating-maternal-mortality-save-lives-south-sudan

14. Salehi M, Zivkovic I, Mayronne S, Letoquart JP, Joharifard S, Joos E. The Evaluation of a Surgical Task-Sharing Program in South Sudan. Surgeries. 2023;4(2):175–87.

15. Cherongis M. An interview with Dr Lul Lojok Deng, Director General of the Public Health Laboratory, Ministry of Health of the Republic of South Sudan [Internet]. Amref Health Africa Newsroom. 2022. Available from: https://newsroom.amref.org/news/2022/05/an-interview-with-dr-lul-lojok-deng/

16. Bayo P, Itua I, Francis SP, et al. Estimating the met need for emergency obstetric care (EmOC) services in three payams of Torit County, South Sudan. BMJ Open. 2018;8(2):e018739.

17. Macrotrends.net. South Sudan Infant Mortality Rate 1950-2023 [Internet]. 2023. Available from: https://www.macrotrends.net/countries/SSD/south-sudan/infant-mortality-rate

18. City Population. Central Equatoria (State, South Sudan) - Population Statistics, Charts, Map and Location [Internet]. [cited 2024 Jan 28]. Available from: https://www.citypopulation.de/en/southsudan/admin/92central_equatoria/

19. Bruno E, White MC, Baxter LS, et al. An Evaluation of Preparedness, Delivery and Impact of Surgical and Anesthesia Care in Madagascar: A Framework for a National Surgical Plan. World J Surg. 2017;41(5):1218–24.

20. Vyas RM, Sayadi LR, Bendit D, Hamdan US. Using Virtual Augmented Reality to Remotely Proctor Overseas Surgical Outreach: Building Long-Term International Capacity and Sustainability. Plast Reconstr Surg. 2020;146(5):622e–9e.

21. World Medical Association. World Medical Association Declaration of Helsinki: ethical principles for medical research involving human subjects. JAMA. 2013;310(20):2191–4.

22. Debas HT, Donkor P, Gawande A, et al., editors. Essential Surgery. Disease Control Priorities, 3rd edition, Volume 1. Washington (DC): The International Bank for Reconstruction and Development / The World Bank; 2015.

